# The Statistical Illusion of a Global HIV Endgame

**DOI:** 10.1101/2025.03.30.25324894

**Authors:** Shaoming Chen

## Abstract

This study analyzes prevalence growth rates across regions using UNAIDS data and statistical modeling to forecast trends through 2050. Globally, the 95-95-95 targets are projected to be achieved by 2035, 2034, and 2025, respectively, largely due to the success of robust interventions in Eastern and Southern Africa. However, this masks the continued high burden and rapid accumulation of new infections in Asia and the Pacific, the Caribbean, and the Middle East and North Africa, where persistent transmission and limited treatment coverage reveal stark disparities. These regional crises challenge the framework’s global applicability, suggesting Africa’s progress cannot be extrapolated universally. Relying solely on the 95-95-95 framework risks creating an illusory sense of achieving an ‘endgame’ without tailored strategies that address diverse epidemiological contexts. This study used a random forest model analyzing feature correlations with HIV growth. The new infection is most strongly correlated, while the first 95 target is also highly correlated, indicating that prevention and high testing significantly reduces the growth rate. The LGBT score and urban population are positive correlated, suggesting that HIV growth is primarily driven by the LGBT community in highly urbanized areas. The third 95 target displays weaker correlations, showing that HIV growth is driven by those who have not been tested. Prevention efforts should prioritize encouraging untested individuals to undergo testing in the urban LGBT populations.

## Background

UNAIDS, a key agency of the United Nations, has established the 95-95-95 Targets to end the AIDS epidemic by 2030 ^[1]^, aligning with global health objectives. These targets aim to reduce new HIV infections and AIDS-related deaths while ensuring that the majority of people living with HIV have access to life-saving treatment. Although significant progress has been achieved in some areas, the global response remains uneven, and multiple factors continue to hinder these efforts.

One notable achievement has been a 60% reduction in new HIV infections since 1995 ^[2]^, but many regions are falling behind on key targets. In Asia and the Pacific, for example, only 79% of people living with HIV know their status, which is far from the 95% target. Globally, 9 million people still lack access to treatment, increasing their risk of poor health outcomes. ^[3]^ Beyond economic factors, challenges such as inadequate healthcare infrastructure, political instability, social stigma, and limited access to education and prevention programs significantly influence the effectiveness of HIV interventions ^[4]^.

This research will analyze current data on new infections and related factors to project future trends.

By examining these factors, the study aims to identify gaps in the response and offer actionable insights to improve intervention strategies.

## Methods

This study employs a comprehensive approach to analyze trends in HIV/AIDS and forecast future trajectories through 2050, utilizing global health and demographic data. The methodology integrates quantitative data analysis and statistical modeling to evaluate progress toward the UNAIDS 2030 goals and predict long-term outcomes.

The primary data source for this research is the UNAIDS HIV estimates with uncertainty bounds, covering the period from 1990 to the present ^[4]^. These documents provide detailed statistics on HIV prevalence, incidence, testing, treatment, and mortality, along with uncertainty ranges to account for variability in reporting. The analysis begins with an examination of global trends to establish an overall picture of the epidemic’s evolution. Subsequently, the data are disaggregated to explore regional patterns, followed by a detailed investigation of individual countries where sufficient data are available. Countries with no data are excluded from the analysis.

Python is used as the primary programming language for data analysis, offering robust tools for handling large datasets and performing statistical modeling. Specifically, the Pandas ^[5]^ library is employed to process Excel tables from UNAIDS and the United Nations, enabling data cleaning, aggregation, and transformation into a suitable format for analysis. The mathematical models are applied to fit the historical data and project future trends. These models are selected to capture different potential trajectories of the epidemic, representing linear growth, or slowing progression, respectively. The forecasting formulas for each model are outlined in **Table 1** below, providing a structured basis for extrapolation beyond the current data.

**Table 1.**
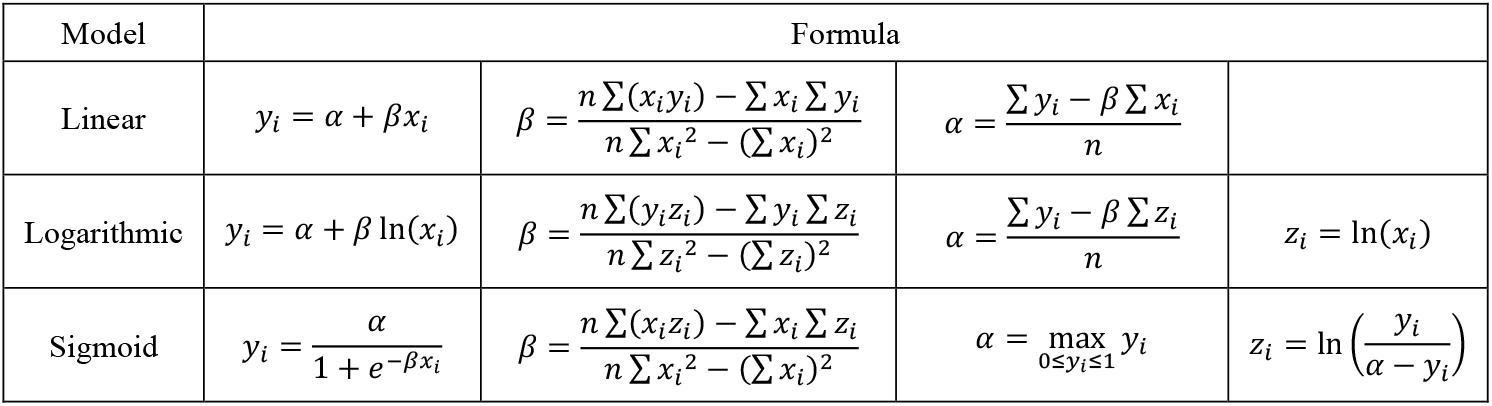
Mathematical models.

| Model | Formula |  |  |  |
| --- | --- | --- | --- | --- |
| Linear | $y_i = \alpha + \beta x_i$ | $\beta = \frac{n \sum(x_i y_i) - \sum x_i \sum y_i}{n \sum x_i^2 - (\sum x_i)^2}$ | $\alpha = \frac{\sum y_i - \beta \sum x_i}{n}$ | |
| Logarithmic | $y_i = \alpha + \beta \ln(x_i)$ | $\beta = \frac{n \sum(y_i z_i) - \sum y_i \sum z_i}{n \sum z_i^2 - (\sum z_i)^2}$ | $\alpha = \frac{\sum y_i - \beta \sum z_i}{n}$ | $z_i = \ln(x_i)$ |
| Sigmoid | $y_i = \frac{\alpha}{1 + e^{-\beta x_i}}$ | $\beta = \frac{n \sum(x_i z_i) - \sum x_i \sum z_i}{n \sum x_i^2 - (\sum x_i)^2}$ | $\alpha = \max_{0 \leq y_i \leq 1} y_i$ | $z_i = \ln\left(\frac{y_i}{\alpha - y_i}\right)$ |

For the curve-fitting process, the NumPy ^[6]^ and SciPy ^[7]^ libraries are used, with the *curve_fit* function from SciPy enabling precise parameter estimation for each model. The fitted models are then used to forecast HIV-related metrics, such as new infections, treatment coverage, and viral suppression, up to the year 2050.

To assess the accuracy of the fitted models, the coefficient of determination *R*^2^ is calculated for each model, providing a measure of how well it explains the variance in the historical data. *R*^2^ values are computed using the *r*2*score* function from the Scikit-learn ^[8]^ library, ensuring a standardized and reliable evaluation metric. The model with the highest *R*^2^ is selected as the best fit for that specific dataset, indicating the most accurate representation of the observed trends and the most reliable basis for forecasting.

This study used a random forest regression method to predict HIV growth rate, with features including LGBT score ^[9]^, sex education ^[10]^, urban population ^[11]^, funding ^[12]^, and three indicators of the 95-95-95 target ^[3]^. Due to limitations in data sources, quantitative indicators of key variables such as social stigma and legal barriers were not included. Random forest was chosen because of its robustness to heterogeneous datasets and its ability to handle high-dimensional features. Random forest is an ensemble learning method that performs well in dealing with complex, nonlinear, multidimensional data by building multiple decision trees and aggregating their predictions. ^[13]^ This study set estimators to 100 and random state to 1 to ensure the stability and repeatability of the model. Feature importance is calculated by averaging the feature contribution of each tree, providing the relative impact of each variable on the prediction, while the Pearson correlation coefficient is used to reveal the positive or negative relationship between the feature and the target variable.

## Results

This analysis utilizes data from UNAIDS spanning 2013 to 2024 to fit the growth trend of the global HIV epidemic. Based on this projection, the first, second, and third targets of the 95-95-95 framework—95% of people living with HIV knowing their status, 95% of diagnosed individuals receiving treatment, and 95% of those treated achieving viral suppression are estimated to be achieved around 2035, 2034, and 2025, respectively. These trends are illustrated in **Figure 1** below.

**Figure 1.**
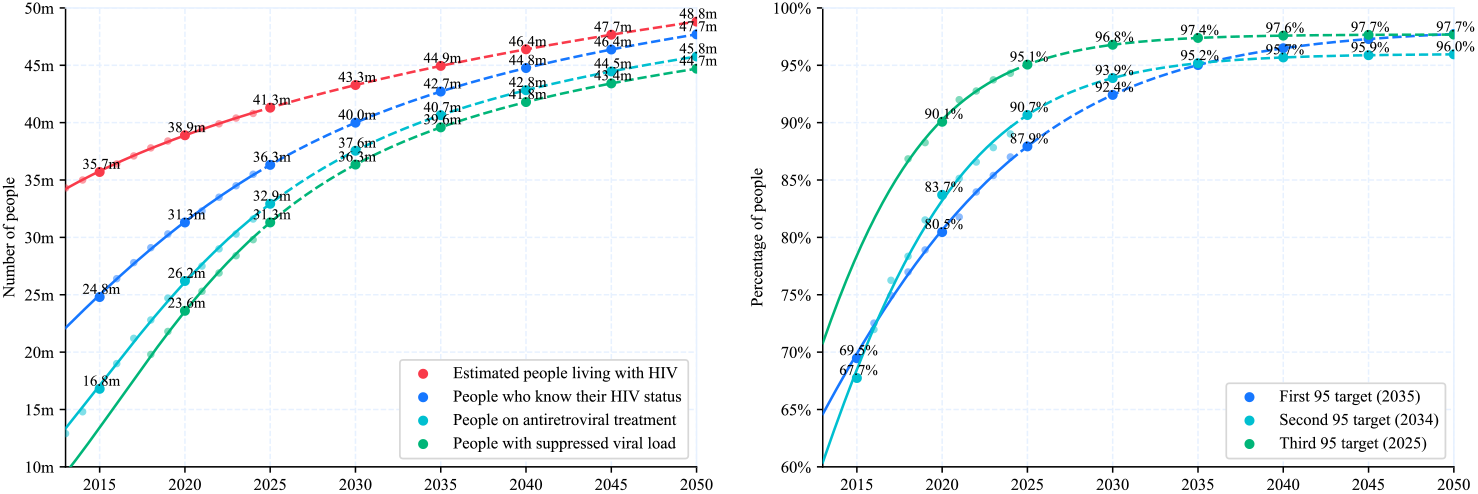
Projected timeline for achieving the UNAIDS 95-95-95 targets based on global HIV data from 2013 to 2024. The figure illustrates the estimated years of attainment for the first (95% awareness of HIV status), second (95% treatment coverage), and third (95% viral suppression) targets, projected to occur around 2035, 2034, and 2025, respectively.

However, when zooming in to specific regions, the conclusions diverge significantly. Eastern and southern Africa, the most populous region, accounted for 20.5 million people living with HIV in 2020, representing 52.7% of the global total of 38.9 million. The slowdown in this region has artificially elevated the global average, masking underlying disparities.

While Eastern and southern Africa and Western and central Africa exhibit a decelerating growth trend, other regions present a contrasting picture, as shown in the **Figure 2**. Asia and the Pacific, depicted in the upper-right panel, continue to experience rapid accumulation, rising from 6.1 million in 2015 to a projected 9.1 million by 2050. Similarly, the Caribbean and the Middle East and North Africa, shown in the lower-right panel, show steep increases, with the Caribbean projected to reach 486,500 cases by 2050 and the Middle East and North Africa nearing 461,400 cases. Eastern Europe and central Asia, as well as Latin America, also raise concerns, with their growth trajectories in the lower-left panel suggesting persistent challenges, reaching 4.1 million and 3.2 million respectively by 2050.

**Figure 2.**
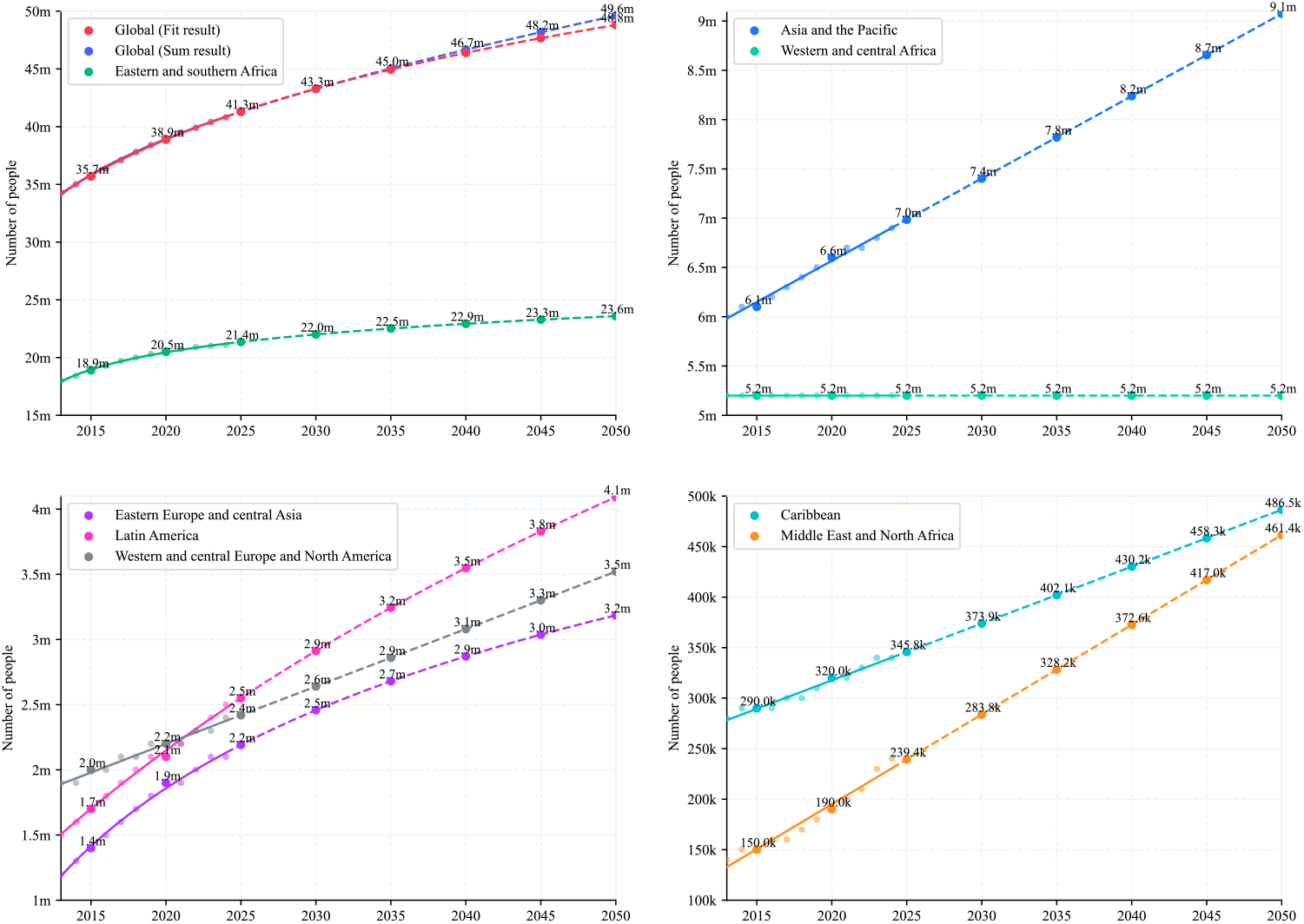
The figure illustrates the projected growth of HIV cases across eight distinct regions, including Eastern and southern Africa, Western and central Africa, Western and central Europe and North America, Eastern Europe and central Asia, Latin America, Asia and the Pacific, the Caribbean, and the Middle East and North Africa. The blue curve indicating the sum of individually fitted regional projections, while the red curve shows the overall fitted global trend. The noticeable divergence between these two curves highlights the disparities masked by averaging the regional data.

The red curve represents the global data, but unlike the previous curve fitting approach, this analysis calculates the actual global growth by summing the projections derived from fitting the data of eight distinct regions individually. A noticeable discrepancy emerges between the summed result and the fitted global trend, with the summed projection reaching 49.6 million by 2050 compared to the fitted global estimate of 48.8 million in 2050, a contrast that is illustrated exclusively in the upper-left panel of **Figure 2**. This gap underscores how averaging obscures the true growth trajectory of the HIV epidemic.

This discrepancy arises because averaging smooths out regional variations, particularly the rapid accumulation in regions like Asia and the Pacific, the Caribbean, and the Middle East and North Africa, while underrepresenting their contribution to the global total. By aggregating data across regions with divergent epidemiological dynamics, such as the decelerating trends in Eastern and southern Africa versus the accelerating trends in Asia and the Pacific, averaging introduces a statistical bias that dilutes the impact of rapid accumulation areas. This methodological flaw highlights the limitations of relying on global averages to capture the heterogeneous nature of the HIV pandemic, potentially leading to over-optimistic conclusions about the trajectory of the epidemic.

For instance, according to UNAIDS statistics, the number of adults and children newly infected with HIV in Asia and the Pacific was 340,000 in 2013, yet by 2024, this figure remained alarmingly high at 300,000. ^[3]^ This marginal decline of only 40,000 over a decade highlights the persistent challenge of controlling new infections in the region, despite global efforts to curb the HIV epidemic. It underscores the limitations of current prevention strategies in addressing the unique social, cultural, and structural barriers in Asia and the Pacific, such as stigma, limited access to testing, and inadequate targeting of high-risk populations, indicating that the region remains a critical hotspot for the ongoing HIV crisis.

The analysis includes data on the LGBT score (Equaldex, 166 countries), sex education (WHO, 117 countries), urban population (World Bank, 163 countries), funding (UNAIDS, 111 countries), new infection and the three indicators of the 95-95-95 target (UNAIDS, 169 countries). However, only 89 countries have complete data across all these indicators. Due to incomplete data across countries, the random forest model was applied only to those with complete datasets. Countries with missing data were excluded from the analysis. For more details, see references 3, 9, 10, 11, and 12. The corresponding datasets are available in the Data Availability Statement. Factors such as social stigma and legal barriers were not included in the analysis, as they are challenging to quantify consistently across countries due to differences in cultural context, legal systems, and the lack of standardized, comparable data.

The dataset contains a variety of socioeconomic and public health indicators, and random forest is able to capture the nonlinear interactions between these variables without assuming a specific functional form. In addition, the method is insensitive to noise and outliers, which is particularly important in cross-national HIV data because the data may vary due to statistical methods or reporting quality. However, as a “black box” model, random forest lacks transparency in its internal decision-making process. Although it can efficiently predict and quantify feature importance, it cannot directly provide clear coefficients or explanatory formulas similar to linear regression. These correlations are illustrated in **Figure 3**.

**Figure 3.**
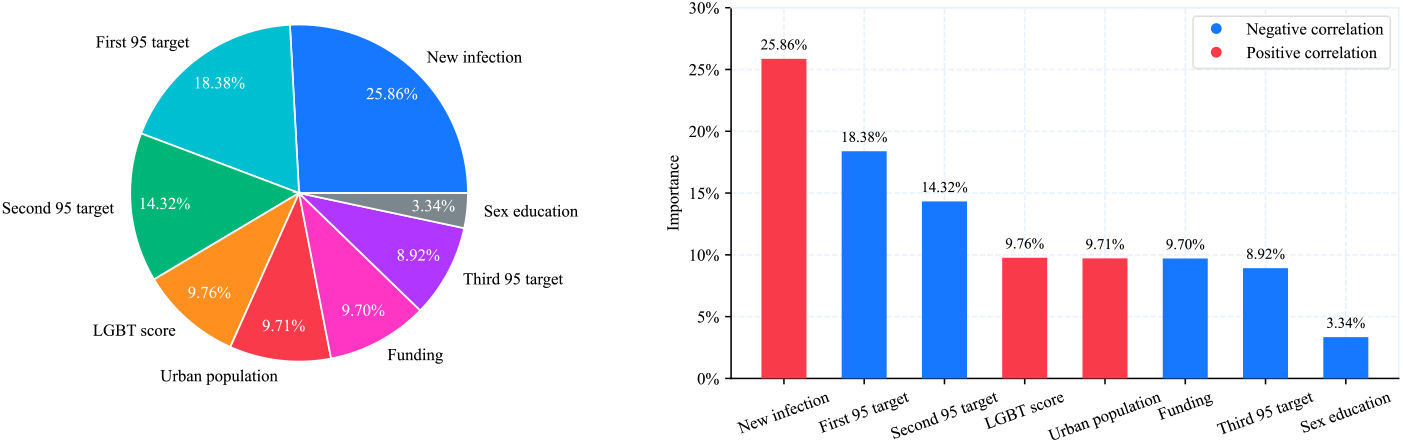
The chart shows a visual analysis of the importance of each feature in the prediction of HIV growth rate. The pie chart on the left uses different colors to represent the relative contribution of each feature, showing its proportion in the model. The bar chart on the right compares the importance of features by bar height, with red and blue marking the positive and negative relationship with the growth rate respectively. The legend distinguishes these two directional influences.

In predicting the HIV growth rate, the correlation between each feature and the growth rate reveals the direction and strength of its impact. The new infection is most strongly correlated, while the first 95 target is also highly correlated, indicating that prevention and high testing significantly reduces the growth rate. The LGBT score and urban population are positive correlated, suggesting that HIV growth is primarily driven by the LGBT community in highly urbanized areas. Funding shows only a general correlation, indicating that its impact is not significant. The third 95 target displays weaker correlations, showing that HIV growth is driven by those who have not been tested. This suggests that the population with viral suppression is not the main factor in virus transmission. As a result, the third 95 target have limited predictive power and contribute minimally to the model.

These findings underscore the importance of prioritizing prevention efforts alongside treatment. While current funding appears to focus primarily on “life-saving activities,” sustained epidemic control also requires proactive strategies to “close the tap” by preventing new infections. Incorporating prevention indicators into epidemic modeling could enhance both statistical robustness and advocacy for upstream interventions.

## Discussion

Relying solely on the proportion of diagnosed people living with HIV (PLHIV), calculated as diagnosed cases divided by the total estimated number of PLHIV, can be misleading when evaluating progress toward HIV testing targets. This indicator, commonly associated with the “first 95” of the UNAIDS 95-95-95 goals, appears to show improvement. For example, in Asia and the Pacific, the proportion increased from 60% in 2017 to 79% in 2024. However, such gains may reflect statistical artifacts rather than substantive progress. If the number of undiagnosed individuals remains relatively constant over time, while the total number of PLHIV increases due to ongoing new infections, estimated at approximately 300,000 annually in the Asia-Pacific region, then the proportion of diagnosed individuals may rise without any actual reduction in the undiagnosed population. Since the numerator reflects the total number of PLHIV minus the undiagnosed population, an increasing denominator combined with a stable number of undiagnosed cases can create the illusion of improved testing coverage.

While achieving the 95-95-95 targets in each subregion is essential for meaningful global epidemic control, it is equally important to consider that the number of new HIV infections has not significantly declined in some contexts. Relying solely on the treatment cascade may obscure ongoing transmission dynamics. Therefore, progress should be assessed not only through the 95-95-95 framework, but also by incorporating additional indicators such as reductions in new infections and improved prevention coverage.

While Eastern and southern Africa have demonstrated significant success, nearing these targets due to robust intervention programs, this progress does not equate to global triumph. The persistent HIV transmission in regions such as Asia and the Pacific, the Caribbean, and the Middle East and North Africa underscores a stark disparity. Relying on the 95-95-95 framework as a marker of global success risks overlooking these regional crises, suggesting that the “endgame” may be an illusion unless tailored strategies address the diverse epidemiological landscapes beyond Africa’s borders.

Additionally, HIV prevention and control strategies should focus on the LGBT community in highly urbanized areas, particularly those who have not been tested. In countries such as China and India, HIV testing coverage among LGBT populations remains low due to factors such as social stigma, limited legal and policy support, and barriers to accessing inclusive healthcare services. The positive correlation between the LGBT score and urban population indicates that these areas have high population density and complex social networks, which may increase the risk of exposure and transmission within the LGBT community. Conversely, individuals who have received treatment, especially those whose virus is suppressed, require less attention. Given limited funding, resources should prioritize encouraging untested individuals to undergo testing.

## Data Availability

All data produced in the present work are contained in the manuscript

https://github.com/rheast/epidemiology

https://www.unaids.org/en/resources/documents/2024/HIV_estimates_with_uncertainty_bounds_1990-present

## Acknowledgments

None.

## Author contributions

Shaoming Chen wrote the manuscript.

## Consent to Publish

Not applicable.

## Consent to Participate

Not applicable.

## Ethics statement

No animals or humans were involved in this study.

## Data availability statement

The datasets analyzed during the current study are available in the GitHub repository at https://github.com/rheast/epidemiology. The data that support the findings of this study are openly available in UNAIDS national HIV estimates files (HIV Estimates with Uncertainty Bounds 1990-Present, 2025, https://www.unaids.org/en/resources/documents/2025/HIV_estimates_with_uncertainty_bounds_1990-present), Equaldex (LGBT Rights by Country & Travel Guide, 2025, https://www.equaldex.com), Joint United Nations Programme on HIV/AIDS (The Journey Towards Comprehensive Sexuality Education: Global Status Report, 2021, https://www.who.int/publications/m/item/9789231004810), World Bank (Urban Population, 2025, https://data.worldbank.org/indicator/SP.URB.TOTL.IN.ZS), and UNAIDS (HIV Financial Dashboard: GARPR 16-GAM 2025 Dataset, 2025, https://hivfinancial.unaids.org/hivfinancialdashboards.html).

